# Females are less likely to receive bystander cardiopulmonary resuscitation in witnessed out-of-hospital cardiac arrest: An Australian perspective

**DOI:** 10.1101/2023.12.19.23300255

**Authors:** Sonali Munot, Janet E Bray, Julie Redfern, Adrian Bauman, Simone Marschner, Christopher Semsarian, Alan Robert Denniss, Andrew Coggins, Paul M Middleton, Garry Jennings, Blake Angell, Saurabh Kumar, Pramesh Kovoor, Matthew Vukasovic, Jason C Bendall, T Evens, Clara K Chow

## Abstract

**Background:** Bystander cardiopulmonary resuscitation (CPR) plays a significant role in survival from out-of-hospital cardiac arrest (OHCA). This study aimed to assess whether bystander CPR differed by patient sex among bystander-witnessed arrests.

**Methods:** Data on all OHCAs attended by New South Wales (NSW) paramedics between January 2017 and December 2019 was obtained from the NSW Public Health Risks and Outcomes Registry (PHROR). This observational study was restricted to bystander-witnessed cases with presumed medical aetiology. OHCA from arrests in aged care, medical facilities, and cases with an advance care directive (do-not-resuscitate) were excluded. Multivariate logistic regression was used to examine the association of patient sex with bystander CPR. Secondary outcomes were OHCA recognition, bystander AED applied, initial shockable rhythm, and survival outcomes.

**Results:** Among the 4,491 bystander-witnessed cases, females were less likely to receive bystander CPR in both private residential (Adjusted Odds ratio [AOR]: 0.82, 95%CI: 0.70-0.95) and public locations (AOR: 0.58, 95%CI:0.39-0.88). Recognition of OHCA in the emergency call was lower for females, particularly in those who arrested in public locations (84.6% vs 91.6%-males, p=0.002) and it partially explained the association of sex with bystander CPR (∼44%). There was no significant difference in OHCA recognition by sex for arrests in private residential locations (p=0.2). Females had lower rates of bystander AED use (4.8% vs 9.6%, p<0.001) however, after adjustment for arrest location and other covariates, this relationship was attenuated and no longer significant (AOR: 0.83, 95%CI: 0.60-1.12). Females were significantly less likely to record an initial shockable rhythm (AOR: 0.52, 95%CI: 0.44-0.61). Although females had greater odds of event survival (AOR: 1.34, 95%CI: 1.15 – 1.56), there was no sex difference in survival to hospital discharge (AOR: 0.96, 95%CI: 0.77-1.19).

**Conclusion:** OHCA recognition and bystander CPR provision differs by patient sex in NSW. Given their importance to patient outcomes, research is needed to understand why this difference occurs and to raise awareness of this issue to the public.

**CLINICAL PERSPECTIVE:** *What is new?:*

- Female OHCA patients in New South Wales, Australia were less likely to receive bystander CPR, irrespective of arrest location.
- In public locations, recognition of OHCA during the emergency call was lower in women and this partly explained the observed sex difference in bystander CPR provision.

*What are the clinical implications?:*

- Public education campaigns and training programs that address bystander response should consider sex differences as a potential barrier to bystander CPR in OHCA
- Future research that examines reasons for lower rates of bystander response in women and ways of addressing this barrier could help address sex disparities in the future.

## INTRODUCTION

Out-of-hospital cardiac arrest (OHCA) is associated with poor survival [1, 2, 3]. Bystander response, including cardiopulmonary resuscitation (CPR) and the use of an automated external defibrillator (AED), is associated with greater survival and better neurological outcomes [4, 5, 6]. However, rates of AED use are suboptimal, and the provision of bystander CPR varies by physical, social, and attitudinal factors related to the bystander and patient [7, 8]. There is also emerging evidence that bystander response may differ depending on the patient’s sex [9, 10, 11].

Lower rates of bystander CPR have been reported for female OHCA patients across several jurisdictions, for example, in the United States and Asia. However, this difference has been reported to vary depending on arrest location, patient age and other factors (e.g., bystander characteristics) [9, 12, 13]. Witnessed status also varies by sex, with females less likely to have a witnessed arrest than men, and this may explain some of the variation seen in bystander CPR [14]. In Australia, a study examining OHCA outcomes noted lower bystander CPR in females, however these were unadjusted estimates [15]. In contrast, a systematic review of data examining sex and OHCA survival, reported higher rates of bystander CPR in female patients [16]. However, their results were based on a comparison of weighted means of bystander CPR percentages and were not adjusted estimates.

Bystander CPR relies on OHCA recognition in the emergency call, which is needed to receive telephone CPR instructions[17]. It has been suggested that OHCA identification and misperceptions about women being in medical distress as a potential barrier in CPR for female patients [18]. To our knowledge, whether there are sex differences in OHCA recognition has not been examined.

The primary aim of this study was to assess if patients’ sex is associated with bystander CPR. The secondary objectives were to examine whether sex was associated with bystander AED application, shockable rhythm, survival outcomes, and recognition of OHCA by emergency call takers as well as explore whether this mediated the association between patient sex and bystander CPR provision.

## METHODS

### Study design and setting

This observational study examined data prospectively collected on all OHCAs attended by NSW Ambulance between January 2017 – December 2019. NSW Ambulance provides emergency medical services (EMS) to all of NSW and NSW Ambulance handles over 1.2 million emergency calls annually. NSW has the highest population of any state in Australia (8,153,000 residents as of 30 June 2022), with over three-quarters living in metropolitan areas [19]. NSW Ambulance call-takers are accredited with the International Academies of Emergency Dispatch (IAED) and use the structured call-taking system Medical Priority Dispatch System ^TM^ (MPDS) [20]. OHCA calls include instructions for CPR and defibrillator retrieval. Ethics approval for this study was obtained from The University of Sydney Ethics Committee (Ref: 2021/017).

### Data source

De-identified data on OHCAs was obtained from the NSW Public Health Risks and Outcomes Registry which is maintained by the NSW Ministry of Health. The registry includes all cases of EMS-attended OHCAs [21, 22], and data is collected, coded, and recorded as per the OHCA Utstein Template [23] [24].

### Inclusion and exclusion criteria

Analyses were restricted to bystander-witnessed cases of arrest due to presumed medical causes that were attended by the EMS. Arrests from external causes (drowning, overdose, trauma), paramedic-witnessed, unwitnessed, with an advance care directive (do-not-resuscitate (DNR) order), arrests occurring in nursing homes, medical centres / GP clinics, police stations, correctional facilities/jails and ambulance stations were excluded from our analysis (Figure 1). Patients with unknown or missing sex data were also excluded.

**Figure 1.**
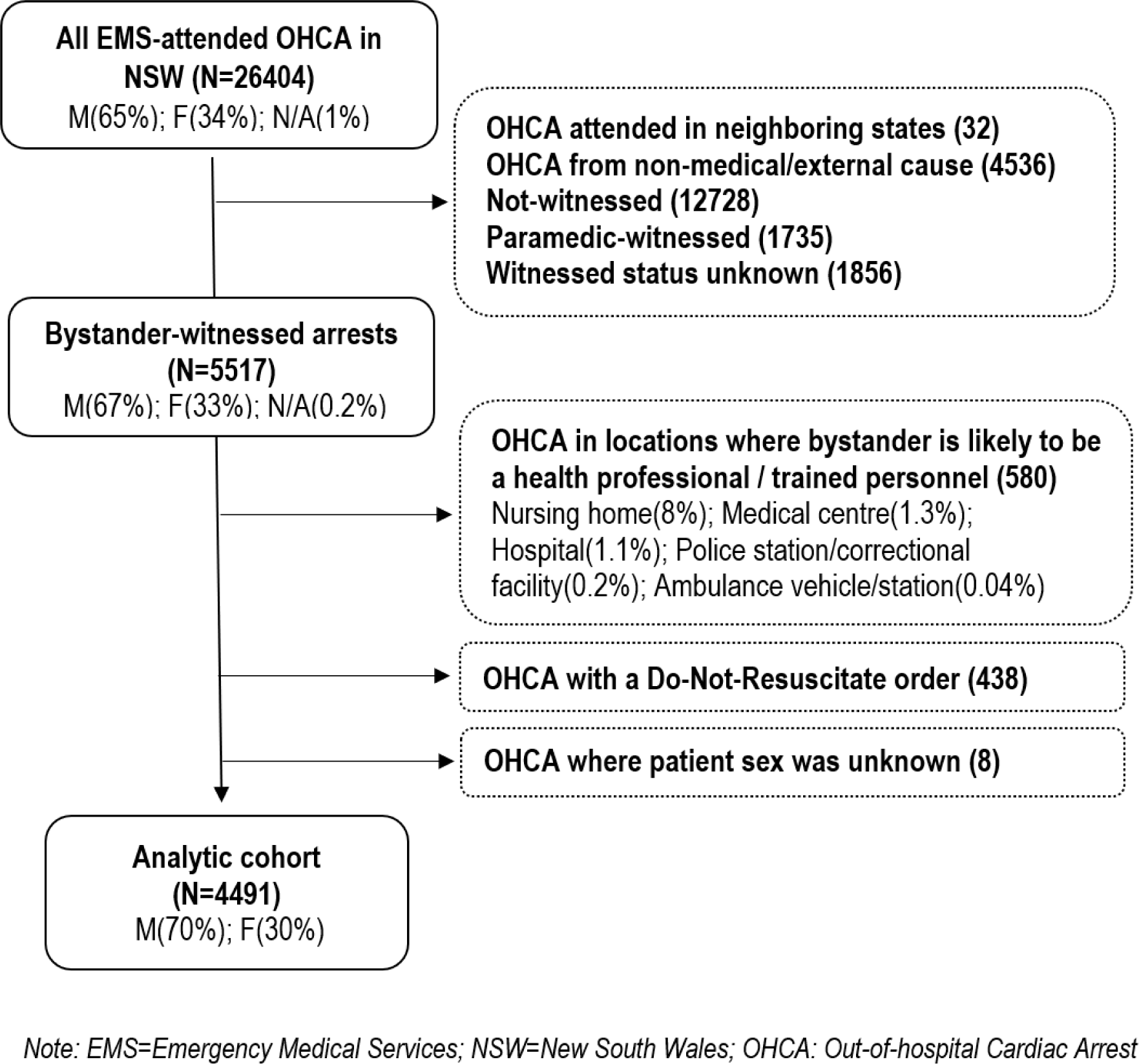
Selection of study analytic cohort.

### Definition of variables

#### Primary and secondary outcome variables

Bystander CPR, defined as “CPR provided by any person who happens to be nearby and is not part of the organised emergency response system”, was categorised as yes or no [21]. A small proportion of responses recorded as unknown/not stated (3.7% females; 2.9% males) were classified as ‘no bystander CPR’ for the purpose of our analyses. Sensitivity analyses were conducted with unknown responses excluded.

Secondary outcomes were: AED application by a bystander (defined as an AED connected to the patient prior to ambulance arrival), initial shockable rhythm, OHCA recognition documented in the emergency call, and, survival outcomes, including event survival (defined as patients with a return of spontaneous circulation on arrival at the hospital emergency department) and survival to hospital discharge.

Recognition of OHCA in the emergency call is documented in the registry as ‘call-taker identified presence of OHCA and is a binary response (yes vs no). Emergency services call-taker assistance is known to influence bystander CPR [7, 25], but requires that the OHCA is recognized in the call. This variable was also assessed as a potential mediator between sex and bystander CPR.

#### Primary independent variable

The primary independent variable was patients’ biological sex, recorded as male or female.

#### Covariates

Patients’ age, arrest aetiology (presumed cardiac vs other medical), witnessed status (yes vs no), arrest location (private residential vs public location), arrest site (urban vs nonurban), advance directive (Do Not Resuscitate order) and ambulance response time were all considered as factors that could potentially influence the association between bystander CPR and patient sex. This was based on previous studies in this space, data availability, and clinical reasoning. These potentially confounding variables were managed by restriction/exclusion, statistical adjustment or presented as subgroups (arrest location: private residential vs public location). Arrest aetiology was collapsed into binary categories of presumed cardiac v/s other medical cause). Non-cardiac medical causes included cancer (6.7%), respiratory disease (6.5%), terminal illness, other (3.0%), neurological (1.3%), Other medical cause, not specified (23.0%). Arrest site areas’ level of remoteness (urban vs nonurban) was defined using the Accessibility and Remoteness Index of Australia (ARIA) score [26]. ARIA classifies an area as urban/metropolitan or nonurban (regional, rural) based on their relative access to services. Missing data for covariates was excluded from analysis (Table 1 footnote).

**Table 1:** Distribution of key arrest/patient characteristics by patient sex in bystander-witnessed OHCA NSW January 2017 – December 2019.

| Characteristic | Study sample<br>N=4491 | Females<br>N=1369 | Males<br>N=3122 | p-value |
| --- | --- | --- | --- | --- |
| **Arrest location** |  |  |  | *<0.001* |
| Private residential | **3366 (75.0)** | 1158 (84.6) | 2208 (70.7) |  |
| Public | **1125 (25.0)** | 211 (15.4) | 914 (29.3) |  |
| **Arrest Site** Urban/metropolitan | **3024 (67.4)** | 933 (68.2) | 2091 (67.1) | 0.50 |
| **Presumed cardiac aetiology** | **2675 (59.6%)** | 744 (54.0%) | 1931 (61.9%) | *<0.001* |
| **Age (years), Median (IQR)** | **69 (58 - 80)** | 71 (59 - 82) | 68 (57 - 78) | *<0.001* |
| **Bystander CPR** | **3186 (71.0)** | 884 (64.6) | 2302 (73.7) | *<0.001* |
| **Bystander AED used** | **366 (8.2)** | 66 (4.8) | 300 (9.6) | *<0.001* |
| **OHCA recognized in call** | **4001 (90.1)** | 1191 (88.5) | 2810 (90.8) | 0.017 |
| **Ambulance response time (minutes), Median (IQR)** | **9 (7 - 14)** | 10 (7, 14) | 9 (7, 13) | 0.06 |
| **Shockable initial rhythm** | **1565 (35.5)** | 301 (22.4) | 1264 (41.2) | *<0.001* |
| **Survived event** | **1270 (28.3)** | 394 (28.8) | 876 (28.1) | 0.60 |
| **Survived to hospital discharge** | **602 (13.8)** | 142 (10.6) | 460 (15.1) | *<0.001* |

### Statistical analysis

Analyses were conducted using R, version 4.1.0 [27]. Descriptive statistics were calculated with categorical data reported as counts and proportions, and continuous data as medians and interquartile range (IQR). Pearson’s ꭓ^2^ test was used to examine group differences and the Wilcoxon rank sum test was used for continuous data. For both tests, the p-value was considered significant if below 0.05. Missing data was excluded from analysis (see Table 1 footnote). Primary analysis involved the examination of the association between patients’ sex and bystander CPR. Multivariate logistic regression models were adjusted for potentially confounding variables that were retained in the model if clinically relevant and associated with bystander CPR at p<0.05 (Table 2). The primary outcome was stratified by arrest location that were grouped into private residential locations (homes) versus public locations.

**Table 2:** Univariate associations of key patient/arrest characteristics and the likelihood of receiving bystander CPR (OR and 95%CI)

|  |  | All arrests | Private residential location | Public location |
| --- | --- | --- | --- | --- |
| **Arrest location** |  |  |  |  |
|  | Public place | Reference |  |  |
|  | Private residential | 0.29 (0.24 - 0.34) | n/a | n/a |
| **Arrest site:** |  |  |  |  |
|  | Urban | Reference | Reference | Reference |
|  | Regional/Rural | 0.86 (0.75 - 0.99) | 0.90 (0.77 - 1.04) | 0.73 (0.51 - 1.05) |
| **Patient sex** |  |  |  |  |
|  | Male | Reference | Reference | Reference |
|  | Female | 0.65 (0.57 - 0.74) | 0.76 (0.66 - 0.88) | 0.59 (0.40 - 0.89) |
| **Patient age** |  |  |  |  |
|  | (per one-year increase in age) | 0.98 (0.97 – 0.98) | 0.98 (0.98 – 0.99) | 0.99 (0.98 – 1.00) |
|  | (By age group) |  |  |  |
|  | <55 years | Reference | Reference | Reference |
|  | 55-75 years | 0.65 (0.53 - 0.78) | 0.71(0.55 - 0.88) | 0.52 (0.32 - 0.83) |
|  | >75 years | 0.35 (0.29 - 0.43) | 0.39 (0.32 - 0.49) | 0.56 (0.32 - 0.96) |
| **Arrest aetiology** |  |  |  |  |
|  | Presumed cardiac | Reference | Reference | Reference |
|  | Other medical | 0.50 (0.44 - 0.58) | 0.52 (0.45 - 0.60) | 0.73 (0.51 - 1.05) |
| **OHCA recognized in call** |  |  |  |  |
|  | Yes | Reference | Reference | Reference |
|  | No | 0.15 (0.12 - 0.18) | 0.13 (0.10 - 0.17) | 0.11 (0.07 - 0.18) |
| **Ambulance response time (minutes)** |  |  |  |  |
|  | <5 | Reference | Reference | Reference |
|  | 5-7 | 1.64 (1.25 – 2.14) | 1.55 (1.13 – 2.14) | 3.65 (1.98 – 6.99) |
|  | 7-10 | 1.50 (1.18 – 1.91) | 1.52 (1.13 – 2.03) | 2.86 (1.71 – 4.80) |
|  | 10-14 | 1.32 (1.03 – 1.68) | 1.38 (1.03 – 1.85) | 2.69 (1.56 – 4.71) |
|  | >14 | 1.10 (0.86 – 1.39) | 1.19 (0.89 – 1.58) | 1.84 (1.10 – 3.07) |

*Note: OR: Odds ratio; CI: Confidence interval; Arrest site (Urban vs nonurban); OHCA: Out-of-hospital cardiac arrest*

Public locations included public building /public place (15.1%), street / road / highway (2.7%), sporting / recreation event (2.3%), vehicle (1.4%), workplace / industrial (1.4%), airport (0.7%), school /educational institution (0.4%), public transport (0.3), other-not specified (0.7%). Multivariable models for secondary outcomes use the total sample and were not split by location, given the limited sample size.

Mediation analysis was conducted to test whether the association between the patients’ sex and bystander CPR could be potentially explained by recognition of OHCA during the emergency call, and this was examined using the *mediation package* in R [28] (Supplementary section 2). This required comparing regression models with and without the proposed mediator variable and involved a bootstrapping approach to arrive at an estimate of the proportion mediated [29, 30]. Mediation was assessed when the following prerequisites were fulfilled: (a) the independent variable (patient sex) affects the mediator (OHCA recognition) (b) the mediating variable affects the outcome (bystander CPR) [31, 32]

## RESULTS

In the three years between January 2017 and December 2019, NSW Ambulance attended 21,836 OHCAs from a medical cause (Figure 1). Of the bystander-witnessed cohort (n=4491), 30% were female (Table 1). Most arrests occurred in private residential locations, although this was significantly higher in females (84.6% females vs 70.7% males, p<0.001) (Table 1). Females were also older (median age: 71 vs 68 years, p<0.001), and less likely to have a presumed cardiac cause than males (54.0% females vs 61.9% males, p<0.001). The majority of bystanders in private residential locations were related to the patient as compared with those in a public location (72.3% vs 6.5%, p<0.001) (Supplementary Table S1). The rate of OHCA recognition documented during the emergency call was significantly lower for females that arrested in a public location (84.6% vs 91.6%, p=0.002), but was not significantly different for arrests in private residential locations (89.2% vs 90.5%, p=0.2) (Supplementary section Table S1). Ambulance response times were similar for male and female patients irrespective of location (p=0.06). Compared with males, bystander CPR was significantly lower for female patients overall (64.6% vs 73,7%, p<0.001), and in both private residential (61.5% vs 67.8%, p<0.001) and public locations (81.5% vs 88.2%, p=0.010) (Supplementary section Table S1).

In sensitivity analysis, excluding cases where bystander CPR status was unknown/not stated did not make a significant difference to the results.

The likelihood of bystander CPR was significantly lower with increasing age (OR: 0.98 95%CI: 0.97 – 0.98); in arrests presumed to be of a non-cardiac medical aetiology (OR: 0.50 95%CI: 0.44 – 0.58); when OHCA was not recognised during the emergency call (OR: 0.15 95%CI: 0.12 – 0.18) and when the ambulance arrived at the arrest scene in under five minutes (Global p<0.0001) (Table 2).

After adjusting for covariates, females had significantly lower odds of receiving bystander CPR (Private location: AOR 0.82, 95%CI:0.70 - 0.95; Public location: AOR 0.58, 95%CI:0.39 - 0.88) (Figure 2). The association between patient sex and bystander CPR in public locations was partially mediated (estimate ∼44%) by recognition of OHCA in the call – with the inclusion of this variable in the adjusted model attenuating the association (between patient sex and bystander CPR) (AOR: 0.67 95%CI:0.43 – 1.06).

**Figure 2:**
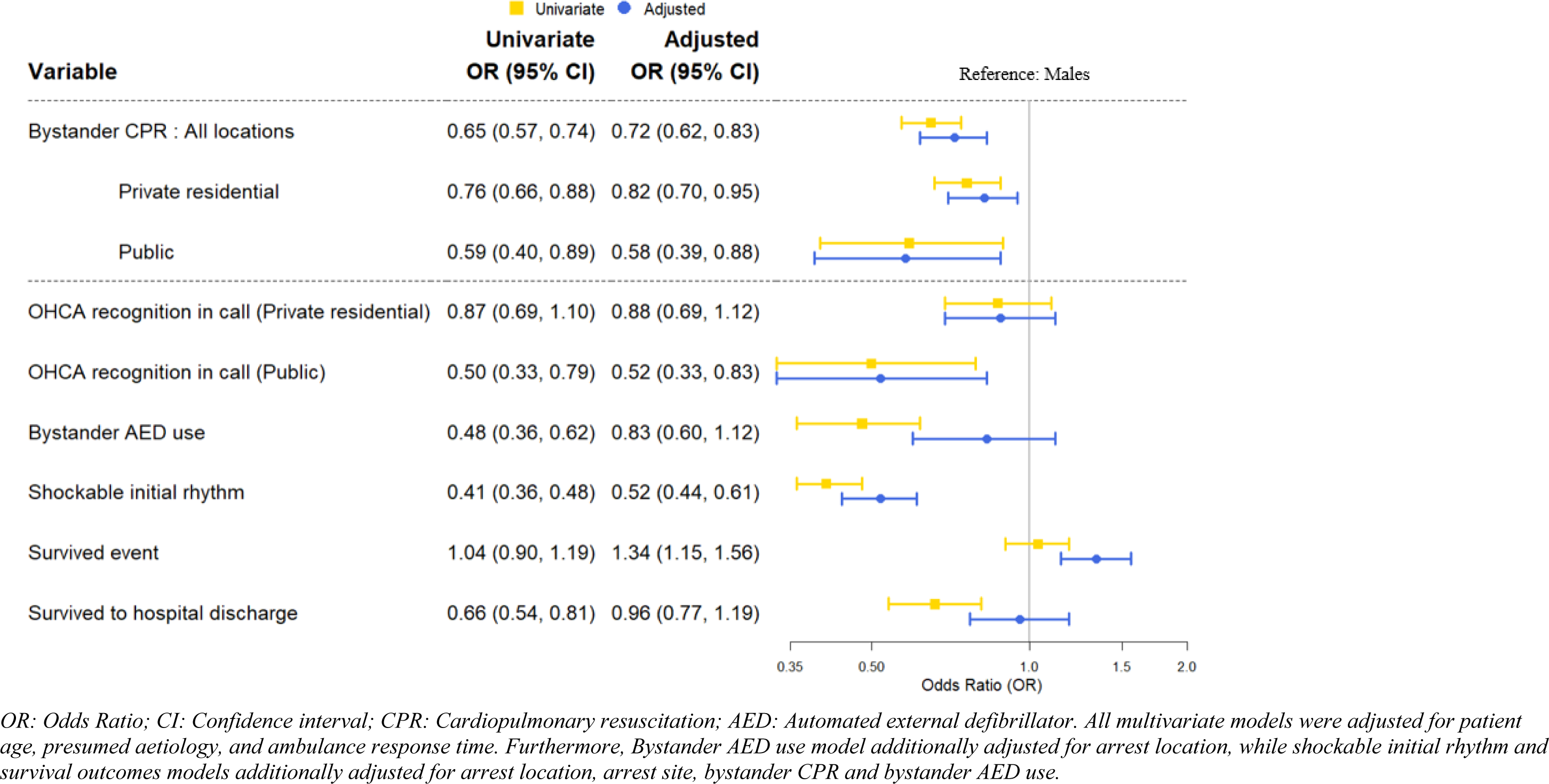
Crude and Adjusted odds ratios explaining the association between patient sex with primary and secondary outcomes.

### Secondary outcomes

OHCA recognition documented by the emergency call taker was lower for females arresting in public locations (AOR: 0.52 95%CI: 0.33 – 0.83). Bystander AED use was significantly lower among females (4.8% vs 9.6% p<0.001) (Table 1). Most of AED application reflects use in public locations compared with private residential locations (28.3% vs 1.4%, p<0.001) (Supplement Table S1). After adjusting for location there was no significant difference in AED application by patient sex (AOR 0.83 95%CI:0.60-1.12) (Figure 2). Females had lower odds of presenting with a shockable initial rhythm compared with male patients (AOR 0.51, 95%CI:0.43 - 0.60) (Figure 2). Females had a greater likelihood of surviving the event to reach the emergency department (AOR: 1.34, 95%CI:1.15 - 1.56), but this survival advantage was not sustained to hospital discharge (AOR: 0.96 95%CI:0.77 - 1.19).

## DISCUSSION

Female OHCA patients are less likely to receive bystander CPR compared with male patients and this association was significant after accounting for covariates including age, presumed aetiology, and ambulance response time. This relationship was consistent for both public locations and private residential locations. Emergency call takers were less likely to document recognition of OHCA in females, and mediation analysis demonstrated that this partially explained the lower rates of bystander CPR in females. Rates of bystander AED use were low overall and while females were less likely to have an AED applied, they were also less likely to present in shockable rhythm, however the association of sex with AED use became non-significant after adjusting for covariates. Survival to hospital discharge was similar by sex.

Several studies have found differences in provision of bystander CPR by sex with most indicating lower rates in females and some suggesting the observed differences vary by arrest location or patient age [9, 11, 12, 14]. For example in the United States, males had higher odds of receiving bystander CPR in public locations, but not in private residential locations[9]. Similar observations were made across Asian countries where an analysis of 56,192 OHCA cases found females had lower rates of bystander CPR in public locations, but in private locations there were higher rates of CPR for females compared to males[11]. Such observations have been explained as potentially due to the bystander knowing or being related to the patient in private locations versus a discomfort of touching the chest of an unknown female in public locations [9, 13, 18]. Indeed, one study form the U.S. suggests it may be less socially acceptable to perform CPR in women with hesitancy in touching females suggested as a factor in a public survey conducted in the United States [18]. In our cohort, majority of bystanders were related with patients in private residential locations, less so in public locations. Despite this we observed a disadvantage in CPR provision for females in private residences.

A variety of factors have been hypothesized to explain lower rates of bystander CPR in women including concerns around modesty, fear of causing harm or legal liability and perceptions of fragility[18, 33]. OHCA may present differently in females compared to males. The current study demonstrating the contributing role of recognition of OHCA by the call taker is also consistent with a mixed methods investigation in which audio recordings of emergency calls were analysed to examine factors associated with emergency call-takers sensitivity in OHCA recognition finding a lower recognition in females [34]. Call-takers generally apply standard algorithms in triage and identification of OHCA [35] which depend on the caller’s description of the patient’s condition. The lower sensitivity in OHCA identification for female could be related to callers’ description of symptoms or the seriousness of patients’ condition. Blom et al (2019) examined if there were delays in OHCA recognition by assessing the time from emergency call to ambulance dispatch but found no difference by patient sex [14]. They noted that they could not factor in delays from OHCA onset to recognition by bystanders. Researchers have pointed to sex-related differences in warning symptoms prior to cardiac arrest noting that while chest pain was more commonly experienced by men, women more typically had shortness of breath [36, 37]. Linguistic factors were also found to be important in influencing whether the emergency call will progress to bystander CPR provision [38, 39]. Future investigations that involve listening to emergency call recordings and analysing the interaction between caller and call-taker may be able to specifically identify barriers unique to women.

Our findings noted that while rates of bystander AED use were lower among females, the difference was not significant after accounting for covariates. Women were significantly more likely to arrest in residential locations compared with males and use of AEDs in private residential locations was very low. Studies from larger populations in the United States and Japan have found that men were significantly more likely to have public AEDs applied by a bystander [10, 40]. They speculated that the differences observed could relate to embarrassment or fear of sexual assault [10, 40].

Females also had a lower likelihood of presenting in a shockable initial rhythm irrespective of age and location. This could be related to differences in arrest aetiology and mechanisms of cardiac arrest [15]. However, a lack of or a delay in CPR provision could also play a role, given that over time shockable rhythms degenerate to non-shockable rhythms without chest compressions [41]. As reported in other studies, females were more likely to survive to hospital, but there was no difference in survival to hospital discharge [15, 42]. Several studies have examined the differences in aetiology and comorbidities among women. However, it is uncertain whether a real difference in survival exists after accounting for known patient, prehospital and treatment factors that could explain disparities [42].

Our study has limitations. We were limited in our ability to control for unmeasured confounders that could explain the observed sex-based disparities (e.g., bystander characteristics, perceived frailty, comorbidities) [43, 44]. We controlled for this to some extent by excluding arrests with a DNR order and nursing home/medical facility arrests where females were overrepresented. Additionally, we adjusted for age and arrest aetiology given the higher age at arrest in females and a greater rate of non-cardiac causes (e.g., terminal illness) compared with males. Witness status was missing for several cases and these cases were excluded from our analysis [14, 45]. The registry data did not distinguish if bystander CPR provision was spontaneous (bystander-initiated) or in-time (telephone guided). Dispatcher assistance has been shown to influence initiation and quality of bystander CPR [46] and rates of recognition by OHCA by emergency call takers were high, suggesting that dispatcher assistance could be high in this cohort [47, 48]. The mediation analysis examining the role of OHCA recognition during the emergency call should only be considered as hypothesis generation of the suggested mechanism rather than definitive evidence of causal processes given that it is based on non-experimental or observational data [32, 49]. Finally, our sample size limited precision and analysis of secondary outcomes and sub-groups.

## CONCLUSION

This study provides novel new data demonstrating in Australia’s most populous state which has high rates of CPR training in the general population, females are less likely to receive bystander CPR in OHCA. It also describes a potential mitigating mechanism for the observation of sex differences with demonstrating the potential role of call takers in recognising OHCA over the phone. The findings suggest that public education, campaigns are needed to address these inequalities and possibly the utility of targeting emergency personnel to help with redressing the issue of recognition of possible OHCA over a call. However further research is needed to better understand this issue and to also develop interventions to address them.

## AUTHOR CONTRIBUTIONS STATEMENT

SM conceived the study design, submitted the ethics application, analysed the data, interpreted the results, and wrote the first draft of this manuscript. JB and SMarschner assisted with analysis. All authors (SMunot, JB, JR, AB, SMarschner, CS, ARD, AC, PMM, GJ, BA, SK, PK, MV, JB, ET, CC) have reviewed, critiqued, and provided intellectual input on various drafts and approved the submitted draft. CC acquired the data, critiqued the analysis plan, multiple drafts and approved the final draft and is the overall guarantor.

## FUNDING AND ACKNOWLEDGEMENTS

This work was supported by the National Health and Medical Research Council (NHMRC) of Australia partnership project grant (#1168950). In addition, and as part of the NHMRC partnership grant, the study received support from the following partner organisations: NSW Ministry of Health, Surf Life Saving NSW, Western Sydney Local Health District, NSW Ambulance, The National Heart Foundation of Australia, Michael Hughes Foundation (now merged with Heart of the Nation), Heart Support Australia, City of Parramatta, Take Heart Australia, and the NSW Data Analytics Centre. SMunot was funded by PhD scholarships from The University of Sydney centres (Westmead Applied Research Centre and Charles Perkins Centre Westmead node), JB is funded by a Heart Foundation of Australia Fellowship (##104751), JR is funded by an NHMRC Investigator Grant (GNT1143538), CS is funded by an NHMRC Practitioner Fellowship (#1154992) and NSW Health, BA is supported by an NHMRC Emerging Leadership Grant (GNT2010055). The authors would like to acknowledge the data custodian NSW Ministry of Health and the statistical assistance of Haeri Min and Desi Quintans.

## CONFLICT OF INTEREST

None

## Data Availability

Data is confidential and we do not have sharing permissions.

## Notes

### Competing Interest Statement

The authors have declared no competing interest.

### Clinical Trial

n/a

### Author Declarations

The University of Sydney Human Research Ethics Committee Ref. no 2021/017.

## REFERENCES

1. Bray J, Howell S, Ball S, Doan T, Bosley E, Smith K, et al. The epidemiology of out-of-hospital cardiac arrest in Australia and New Zealand: A binational report from the Australasian Resuscitation Outcomes Consortium (Aus-ROC). Resuscitation. 2022 Mar;172:74–83.

2. Gräsner JT, Wnent J, Herlitz J, Perkins GD, Lefering R, Tjelmeland I, et al. Survival after out-of-hospital cardiac arrest in Europe - Results of the EuReCa Two study. Resuscitation. 2020;148:218–226.

3. Cardiac arrest registry to enhance survival (CARES). Annual report 2022. Available from: https://mycares.net/sitepages/data.jsp. accessed on 15 November 2023.

4. Song J, Guo W, Lu X, Kang X, Song Y, Gong D. The effect of bystander cardiopulmonary resuscitation on the survival of out-of-hospital cardiac arrests: A systematic review and meta-analysis. Scand J Trauma Resusc Emerg Med. 2018;26(1):86.

5. Bækgaard JS, Viereck S, Møller TP, Ersbøll AK, Lippert F, Folke F. The effects of public access defibrillation on survival after out-of-hospital cardiac arrest: A systematic review of observational studies. Circulation. 2017;136(10):954–965.

6. Odom E, Nakajima Y, Vellano K, Al-Araji R, Coleman King S, Zhang Z, et al. Trends in EMS-attended out-of-hospital cardiac arrest survival, United States 2015–2019. Resuscitation. 2022;179:88-93.

7. Case R, Cartledge S, Siedenburg J, Smith K, Straney L, Barger B, et al. Identifying barriers to the provision of bystander cardiopulmonary resuscitation (CPR) in high-risk regions: A qualitative review of emergency calls. Resuscitation. 2018;129:43–47.

8. Munot S, Rugel EJ, Von Huben A, Marschner S, Redfern J, Ware S, et al. Out-of-hospital cardiac arrests and bystander response by socioeconomic disadvantage in communities of New South Wales, Australia. Resuscitation Plus. 2022;9:100205.

9. Blewer AL, McGovern SK, Schmicker RH, May S, Morrison LJ, Aufderheide TP, et al. Gender disparities among adult recipients of bystander cardiopulmonary resuscitation in the public. Circulation: Cardiovascular Quality and Outcomes. 2018;11(8):e004710.

10. Jadhav S, Gaddam S. Gender and location disparities in prehospital bystander AED usage. Resuscitation. 2021;158:139–142.

11. Liu N, Ning Y, Ong MEH, Saffari SE, Ryu HH, Kajino K, et al. Gender disparities among adult recipients of layperson bystander cardiopulmonary resuscitation by location of cardiac arrest in Pan-Asian communities: A registry-based study. eClinicalMedicine. 2022;44.

12. Ishii M, Tsujita K, Seki T, Okada M, Kubota K, Matsushita K, et al. Sex- and Age-Based Disparities in Public Access Defibrillation, Bystander Cardiopulmonary Resuscitation, and Neurological Outcome in Cardiac Arrest. JAMA Network Open. 2023;6(7):e2321783–e2321783.

13. Matsuyama T, Okubo M, Kiyohara K, Kiguchi T, Kobayashi D, Nishiyama C, et al. Sex-Based Disparities in Receiving Bystander Cardiopulmonary Resuscitation by Location of Cardiac Arrest in Japan. Mayo Clin Proc. 2019;94(4):577–587.

14. Blom MT, Oving I, Berdowski J, van Valkengoed IGM, Bardai A, Tan HL. Women have lower chances than men to be resuscitated and survive out-of-hospital cardiac arrest. Eur Heart J. 2019;40(47):3824–3834.

15. Bray JE, Stub D, Bernard S, Smith K. Exploring gender differences and the “oestrogen effect” in an Australian out-of-hospital cardiac arrest population. Resuscitation. 2013;84(7):957–963.

16. Lakbar I, Ippolito M, Nassiri A, Delamarre L, Tadger P, Leone M, et al. Sex and out-of-hospital cardiac arrest survival: a systematic review. Ann Intensive Care. 2022;12(1):114.

17. Kurz MC, Bobrow BJ, Buckingham J, Cabanas JG, Eisenberg M, Fromm P, et al. Telecommunicator Cardiopulmonary Resuscitation: A Policy Statement From the American Heart Association. Circulation. 2020;141(12):e686–e700.

18. Perman SM, Shelton SK, Knoepke C, Rappaport K, Matlock DD, Adelgais K, et al. Public Perceptions on Why Women Receive Less Bystander Cardiopulmonary Resuscitation Than Men in Out-of-Hospital Cardiac Arrest. Circulation. 2019;139(8):1060–1068.

19. Australian Bureau of Statistics. National, state and territory population [Internet] 2023. Available from: https://www.abs.gov.au/statistics/people/population/national-state-and-territory-population/latest-release. accessed on August 20 2023.

20. Priority Dispatch Corp. Medical Priority Dispatch System 13 ed. Salt Lake City, Utah, USA 2017.

21. Packham N WS, Faddy SC, Fouche PF, Arnold J, Burns B & Bendall JC. Out-of-Hospital Cardiac Arrest in NSW 2020 Annual Report. Sydney: NSW Ambulance; 2020. Available from: https://www.ambulance.nsw.gov.au/data/assets/pdf_file/0006/643722/DE487-OHCAR-Report-2019_V6.pdf. accessed on 07 Jan 2022.

22. Centre for Health Record Linkage (CHeReL). CHeReL Datasets and Data Dictionaries 2021. Available from: https://www.cherel.org.au/datasets. accessed on 05 Dec 2022.

23. Perkins GD, Jacobs IG, Nadkarni VM, Berg RA, Bhanji F, Biarent D, et al. Cardiac arrest and cardiopulmonary resuscitation outcome reports: update of the Utstein Resuscitation Registry Templates for Out-of-Hospital Cardiac Arrest: a statement for healthcare professionals from a task force of the International Liaison Committee on Resuscitation (American Heart Association, European Resuscitation Council, Australian and New Zealand Council on Resuscitation, Heart and Stroke Foundation of Canada, InterAmerican Heart Foundation, Resuscitation Council of Southern Africa, Resuscitation Council of Asia); and the American Heart Association Emergency Cardiovascular Care Committee and the Council on Cardiopulmonary, Critical Care, Perioperative and Resuscitation. Circulation. 2015;132(13):1286–1300.

24. Dyson S. NSW ambulance cardiac arrest registry. 2017 report. 2020. Available from: https://www.ambulance.nsw.gov.au/about-us/corporate-publications accessed on 15 Sept 2023.

25. Viereck S, Palsgaard Moller T, Kjaer Ersboll A, Folke F, Lippert F. Effect of bystander CPR initiation prior to the emergency call on ROSC and 30day survival-An evaluation of 548 emergency calls. Resuscitation. 2017;111:55–61.

26. Australian Government: Department of Health. 2.3 Accessibility Remoteness Index of Australia (ARIA) Remoteness Area (RA) in *Accessibility Remoteness Index of Australia (ARIA) Review Analysis of Areas of Concern–Final Report*. Canberra 2011.

27. R Core Team. R: A language and environment for statistical computing. R Foundation for Statistical Computing, Vienna, Austria Vienna, Austria. 2021. Available from: https://www.R-project.org/. accessed on 3 May 2021.

28. Tingley D, Yamamoto T, Hirose K, Keele L, Imai K. mediation: R Package for Causal Mediation Analysis. Journal of Statistical Software. 2014;59(5):1–38.

29. Kim B. “Introduction to Mediation Analysis” UVA Library StatLab 2016. Available from: https://library.virginia.edu/data/articles/introduction-to-mediation-analysis. accessed on 1st December 2023.

30. Baron RM, Kenny DA. The Moderator-Mediator Variable Distinction in Social Psychological Research: Conceptual, Strategic, and Statistical Considerations. Journal of personality and social psychology. 1986;51(6):1173–1182.

31. Judd CM, Kenny DA. Process Analysis:Estimating Mediation in Treatment Evaluations. Evaluation Review. 1981;5(5):602–619.

32. Shrout PE, Bolger N. Mediation in experimental and nonexperimental studies: new procedures and recommendations. Psychol Methods. 2002;7(4):422–445.

33. Kramer CE, Wilkins MS, Davies JM, Caird JK, Hallihan GM. Does the sex of a simulated patient affect CPR? Resuscitation. 2015;86:82–87.

34. Watkins CL, Jones SP, Hurley MA, Benedetto V, Price CI, Sutton CJ, et al. Predictors of recognition of out of hospital cardiac arrest by emergency medical services call handlers in England: a mixed methods diagnostic accuracy study. *Scandinavian Journal of Trauma*, Resuscitation and Emergency Medicine. 2021;29(1):7.

35. Perera N, Birnie T, Whiteside A, Ball S, Finn J. “If you miss that first step in the chain of survival, there is no second step”-Emergency ambulance call-takers’ experiences in managing out-of-hospital cardiac arrest calls. PloS one. 2023;18(3):e0279521.

36. Reinier K, Dizon B, Chugh H, Bhanji Z, Seifer M, Sargsyan A, et al. Warning symptoms associated with imminent sudden cardiac arrest: a population-based case-control study with external validation. The Lancet Digital Health. 2023;5(11):e763–e773.

37. Gnesin F, Mills EHA, Jensen B, Møller AL, Zylyftari N, Bøggild H, et al. Symptoms reported in calls to emergency medical services within 24 hours prior to out-of-hospital cardiac arrest. Resuscitation. 2022;181:86–96.

38. Riou M, Ball S, Whiteside A, Bray J, Perkins GD, Smith K, et al. ’We’re going to do CPR’: A linguistic study of the words used to initiate dispatcher-assisted CPR and their association with caller agreement. Resuscitation. 2018a;133:95–100.

39. Riou M, Ball S, Williams TA, Whiteside A, Cameron P, Fatovich DM, et al. ’She’s sort of breathing’: What linguistic factors determine call-taker recognition of agonal breathing in emergency calls for cardiac arrest? Resuscitation. 2018;122:92–98.

40. Kiyohara K, Katayama Y, Kitamura T, Kiguchi T, Matsuyama T, Ishida K, et al. Gender disparities in the application of public-access AED pads among OHCA patients in public locations. Resuscitation. 2020;150:60–64.

41. Cournoyer A, Chauny JM, Paquet J, Potter B, Lamarche Y, de Montigny L, et al. Electrical rhythm degeneration in adults with out-of-hospital cardiac arrest according to the no-flow and bystander low-flow time. Resuscitation. 2021;167:355–361.

42. Kotini-Shah P, Del Rios M, Khosla S, Pugach O, Vellano K, McNally B, et al. Sex differences in outcomes for out-of-hospital cardiac arrest in the United States. Resuscitation. 2021;163:6–13.

43. Lee G, Ro YS, Park JH, Hong KJ, Song KJ, Shin SD. Interaction between bystander sex and patient sex in bystander cardiopulmonary resuscitation for Out-of-Hospital cardiac arrests. Resuscitation. 2023;187:109797.

44. Levinson M, Mills A. Cardiopulmonary resuscitation - time for a change in the paradigm? Medical Journal of Australia. 2014;201(3):152–154.

45. Rob D, Kavalkova P, Smalcova J, Franek O, Smid O, Komarek A, et al. Gender differences and survival after out of hospital cardiac arrest. The American Journal of Emergency Medicine. 2022;55:27–31.

46. Kwak J, Ok Ahn K, Chan PS. Sex difference in the association between type of bystander CPR and clinical outcomes in patients with out of hospital cardiac arrest. Resuscitation Plus. 2023;13:100342.

47. Ko SY, Ahn KO, Do Shin S, Park JH, Lee SY. Effects of telephone-assisted cardiopulmonary resuscitation on the sex disparity in provision of bystander cardiopulmonary resuscitation in public locations. Resuscitation. 2021;164:101–107.

48. Faddy S, Cohen S, Peresson C. Accuracy in the identification of cardiac arrest by emergency medical dispatchers in New South Wales (NSW), Australia. Resuscitation. 2019;142:e104.

49. Nguyen TQ, Schmid I, Stuart EA. Clarifying causal mediation analysis for the applied researcher: Defining effects based on what we want to learn. Psychol Methods. 2020.

